# Modelling of time-to-events in an ambispective study: illustration with the analysis of *ABO* blood groups on venous thrombosis recurrence

**DOI:** 10.1101/2021.11.20.21266583

**Authors:** Gaëlle Munsch, Louisa Goumidi, Astrid van Hylckama Vlieg, Manal Ibrahim-Kosta, Maria Bruzelius, Jean-François Deleuze, Frits R. Rosendaal, Hélène Jacqmin-Gadda, Pierre-Emmanuel Morange, David-Alexandre Trégouët

**Affiliations:** Univ. Bordeaux, Inserm, Bordeaux Population Health Research Center, UMR 1219, F-33000 Bordeaux, France; INSERM UMR_S 1263, Nutrition Obesity and Risk of Thrombosis, Center for CardioVascular and Nutrition research (C2VN), Aix-Marseille University, Marseille 13385, France; Department of Clinical Epidemiology, Leiden University Medical Center, Leiden, Netherlands; Laboratory of Haematology, La Timone Hospital, Marseille 13385, France; Department of Medicine Solna, Karolinska Institute, Stockholm, Sweden; Department of Hematology, Karolinska University Hospital, Stockholm, Sweden; Université Paris-Saclay, CEA, Centre National de Recherche en Génomique Humaine, 91057, Evry, France; Centre d’Etude du Polymorphisme Humain, Fondation Jean Dausset, Paris, France

**Keywords:** Ambispective, Venous thrombosis, Survival analysis, Recurrent events, Genetics, *ABO* blood groups

## Abstract

In studies of time-to-events, it is common to collect information about events that occurred before the inclusion in a prospective cohort. In an ambispective design, when the risk factors studied are independent of time, including both pre- and post-inclusion events in the analyses increases the statistical power but may lead to a selection bias. To avoid such a bias, we propose a survival analysis weighted by the inverse of the survival probability at the time of data collection about the events.

This method is applied to the study of the association of ABO blood groups with the risk of venous thromboembolism (VT) recurrence in the MARTHA and MEGA cohorts. The former relying on an ambispective design and the latter on a standard prospective one. In the combined sample totalling 2,752 patients including 993 recurrences, compared with the O1 group, A1 has an increased risk (Hazard Ratio (HR) of 1.18, p=4.2×10^−3^), homogeneously in MARTHA and in MEGA. The same trend (HR=1.19, p=0.06) was observed for the less frequent A2 group.

In conclusion, this work clarified the association of *ABO* blood groups with the risk of VT recurrence. Besides, the methodology proposed here to analyse time-independent risk factors of events in an ambispective design has an immediate field of application in the context of genome wide association studies.

## INTRODUCTION

Venous Thrombosis (VT) is a common cardiovascular disease with an annual incidence of ∼1 to 3 per 1,000 in the general population which increases with age.[1] This pathology can manifest as either deep vein thrombosis (DVT) or pulmonary embolism (PE) with a mortality rate within a month of diagnosis at 6% and 12%, respectively.[2]

After a first VT, the recurrence rate is approximately 30% within 10 years.[3] VT recurrence could be prevented by a continued anticoagulant treatment but this therapy leads to a substantial risk of bleeding and a significant cost to society.[4] Understanding the pathophysiological mechanisms of VT recurrence may facilitate the identification of groups of patients at lower risk of recurrence who do not require these treatments. While about 30 loci are now well established to be associated with the genetic susceptibility to VT [5,6], less is known about the genetic susceptibility to VT recurrence which likely differs from that of first VT.[7] Among VT disease loci, the *ABO* locus, coding for the *ABO* blood groups, is one of the most important due to the magnitude of the genetic effects associated with the A1 and B at-risk blood groups and their prevalence in the general population.[8–10] To date, few studies have explored the effect of *ABO* blood types on VT recurrence risk.[11–14] These analyses have generally been conducted in studies of moderate size with few recurrent events and have often relied on serological measurement of blood groups. Recently, our group showed that molecularly defined blood groups are more reliable than serological measurements.[15]

In this work, we wish to investigate the effect of molecularly defined *ABO* blood groups on the risk of recurrence in two large VT cohorts, the MARTHA and MEGA studies.[16,17] In MEGA, participants were included at the time of their first VT, the beginning of the at risk period for the recurrence, and standard time-to-event analysis among which the Cox model is the most popular one[18] can be used for studying the risk of first recurrence. The MARTHA study has a different design since it included all subjects who visited a Thrombophilia centre in Marseille (France) between 1994 and 2012 and had an history of VT (possibly many years before inclusion). Information on recurrence post-inclusion were collected at a follow-up visit several years later but many participants have already experienced a VT recurrence at the time of inclusion. The standard practice is to consider only the information about events that occurred post-inclusion, while excluding patients that have experienced the event of interest (VT recurrence) before their inclusion.[19,20] In the framework of recurrent events, it has also been proposed to analyse all recurrences that fall within the observation window after the inclusion and to stratify the analyses according to the number of events before inclusion.[21] Others have proposed to study only the occurrence of a recurrence during the observation time without excluding patients with a recurrence prior to inclusion and without differentiating between them.[22] In that case, the analysis focused on the association between risk factors measured at inclusion and the risk of recurrence (not necessarily the first one) during the observation time window.

When the risk factors are time-dependent variables such as biological measurements, the analysis including only the events occurring after the collection of the studied variables is needed as the exposure must be measured before the event occurrence in order to avoid bias due to reverse causality. However, when the explanatory variables do not change over time, as genetic factors, this bias is avoided. When the dates of the events that occurred before the inclusion in the study are known, considering this information in the analysis could greatly increase the statistical power. However, specific data analysis procedures should be considered to avoid selection bias by death.

In order to be able to efficiently analyse the impact of *ABO* blood groups on first VT recurrence in MARTHA, a weighted survival analysis is here proposed to enable the joint analysis of patients with or without recurrence prior inclusion in the study.

## MATERIALS AND METHODS

### MARTHA study

This work was motivated by the identification of genetic risk factors for VT recurrence in the MARseille THrombosis Association (MARTHA) study.[23,24] MARTHA includes 2,837 unrelated VT patients who had a consultation visit at the Thrombophilia centre of La Timone Hospital in Marseille (France) between 1994 and 2012. The inclusion date of patients refers to this visit. All patients with at least one documented VT and free of any chronic conditions and of any well characterized genetic risk factors including homozygosity for FV Leiden or Factor II 20210A, protein C, protein S and antithrombin deficiencies, and lupus anticoagulant, were eligible. As an ancillary genetic study, a subsample of 1,592 MARTHA patients have been typed by a high density genotyping arrays, referred thereafter as the MARTHA GWAS subsample (where GWAS stands for Genome Wide Association Study).[25] The MARTHA GWAS sub-study was further extended over the 2013-2018 period and patients were re-contacted to gather information on post-inclusion VT events.

### MARTHA GWAS sub-study and VT recurrence

The previous application of standard quality controls procedures on the genome wide genotype data of MARTHA participants has led to the selection of 1,542 VT patients for genetic analyses.[25] From these remaining individuals, we further excluded patients with autoimmune disease or cancer at inclusion, or with missing information on time to VT recurrence for concerned patients. Finally, 1,518 VT patients were left for the VT recurrence analysis. Among these patients, 411 already had at least one VT recurrence before inclusion. The dates, types (DVT or PE) and provoked characters of the first VT and first recurrence were collected. During the 2013-2018 period, patients were re-contacted to gather information on post-inclusion VT. Among the 1,107 patients with a unique VT at inclusion, 846 (76%) could be re-contacted which led to the identification of 160 additional first recurrences. At the end of the second phase, information on the vital status of non-responders was obtained either through the Répertoire National d’Identification des Personnes Physiques (RNIPP) or medical data. Vital status was finally available for 1,380 individuals including 73 deaths.

### Ambispective design

As the current project aims to assess the effect of *ABO* polymorphisms on the risk of recurrence after a first VT, 4 different types of MARTHA participants can be distinguished (Fig. 1). Case 1 corresponds to patients who had a single VT before inclusion in the study and who were followed up during the recontact phase within which no recurrent event was observed. Case 2 represents patients who had a single VT before inclusion and experienced a recurrent event which was collected during the recontact phase. Case 3 corresponds to patients with a single VT at inclusion for whom no follow-up information was collected during the recontact phase (i.e lost to follow-up). Finally, Case 4 represents patients who had both a first VT and a recurrence before the inclusion in the study. For each of these 4 situations, the at-risk period, a key element in the analysis of recurrent data that represents the period of time that contributes to the estimation of recurrence risk, is shown in grey in Fig. 1.

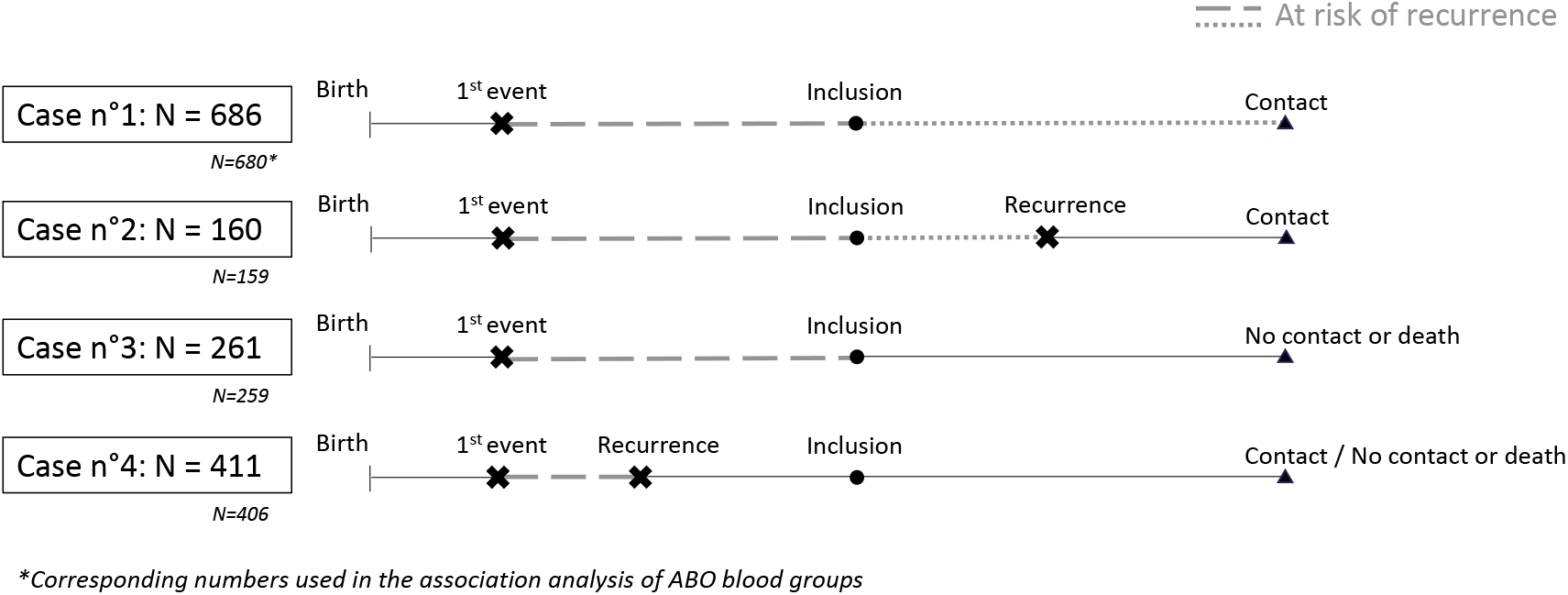

In a standard cohort analysis, only the post-inclusion period of patients from Cases 1&2 (represented with dotted lines), thereafter referred to as the “*prospective sample*”, would be used to investigate risk factors for recurrence. However, since genetic polymorphisms are fixed at birth, all cases of patients can contribute to the analysis of the genetic susceptibility of VT recurrence, considering the first VT as the starting point of the analysis (non-solid lines). This last comment also holds for non-genetic variables available at the time of first VT that are fixed over time such as sex and age at first VT. In the following, we will refer to the “*ambispective sample*” [26] when the four situations are simultaneously considered as it includes both pre- and post-inclusion VT recurrences, that is recurrences which occurred before or within the observation window.

Finally, the MARTHA *prospective sample* was composed of 846 patients including 160 VT recurrences and the extended *ambispective sample* involved 1,518 patients including 571 recurrent VT.

### Statistical modelling of recurrent events using weighted Cox model

The Cox proportional-hazards model is a popular semi-parametric model proposed by Cox in 1972.[27] The relationship between the instantaneous risk function (or hazard function) associated with the occurrence of an event and the vector *Z* of explanatory variables can be written as follows: *λ*(*t, Z, β*) = *λ*_0_(*t*)exp (*β*^T^*Z*) where *β* is the vector of regression coefficients and *λ*_0_(*t*) represents the baseline hazard function.

In order to account for a possible selection bias due to mortality induced by the selection of MARTHA participants, we are proposing a weighted Cox analysis with weights defined by the inverse of the survival probability of individuals up to the time when the information on their possible recurrent event was collected. These weights are used to assign a higher weight to individuals who were less likely to be observed, e.g. individuals at high risk of death before collection of information on VT recurrence.[28] To estimate these weights, we had to model the risk of death in the MARTHA population of VT survivors, using the information on the vital status available for 1,380 patients. As age is the main risk factor for death, we estimated a delayed-entry Cox model with age as time-scale allowing non-parametric modelling of the age effect. For this analysis, subjects contributed from their age of inclusion in the study to their death or age at last information on the vital status. Estimated parameters from this model were used to compute for all MARTHA patients their individual survival probabilities up to the appropriate time point according to their own clinical and covariates information. While for Cases 1&2 patients, the collection of information on VT recurrence is conditional to the survival of patients up to the recontact date, the collection of this information for Case 3&4 patients is conditional to their survival up to their inclusion in the study.

Once these weights are computed, they can be used in a weighted Cox model for the risk of first recurrence, with delay since the first VT as time scale, to analyse both *prospective* and *ambispective* samples. As the prospective analysis considers only post-inclusion events, a model with delayed-entry at the time of inclusion was estimated. This is not necessary for the *ambispective* analysis as all available information since the first VT is then considered. Once Hazard Ratio (HR) association parameters are obtained, their variance can be estimated using the robust method accounting for the within-subject correlation induced by the weights.[29] The weights were computed and standardized using the survival probabilities so that their sum corresponds to the studied sample size with the following formula:

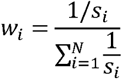

With *w*_*i*_ the weight of *i*^th^ individual, *s*_*i*_ the survival probability of the *i*^th^ individual at its own data collection time and *N* the studied sample size. This approach is implemented with the *survival* package of the R version 3.6.1 environment.[30,31]

### The MEGA study

Briefly, MEGA is a case-control study for VT that includes almost 4,900 patients who were included for their first VT between 1999 and 2004.[32] Among them, 1,289 VT cases had available genetic data. Between 2008 and 2009, questionnaires were sent to patients to gather information on a possible VT recurrence. From the 1,289 VT patients, we excluded 9 individuals who died before the re-contact phase, 17 individuals with missing information on the provoked character of the first VT event and 9 individuals who were homozygous for the factor V Leiden in order to match to MARTHA exclusion criteria. Six patients from the MEGA study (0.5%) were further excluded as it was not possible to unambiguously determine their *ABO* blood group (see next paragraph). Finally, 1,248 MEGA patients including 428 recurrences were included in the analysis. As these patients were included for their first event, a Cox model in which the delay since the first event was employed as time scale to investigate risk factors of first VT recurrence.

### ABO blood groups genetic determination

Five *ABO* polymorphisms were investigated in order to infer *ABO* blood groups. Following recent recommendations[15], the rs8176719-delG was used to tag for O1, the rs41302905-T allele for O2, the rs2519093-T for A1, the rs1053878-A for A2 and the rs8176743-T allele for B.

As MARTHA and MEGA participants have been typed by high-throughput genotyping arrays and imputed on the 1000G Phase I Integrated Release Version 2 Haplotypes, we used best-guessed genotypes from imputed data to infer *ABO* blood groups.[25,33] Note that all 5 polymorphims have imputation quality greater than 0.9 in MARTHA and in MEGA. It was possible to infer *ABO* haplotypes and pair of haplotypes without ambiguity for 1,504 (99.1%) and 1,248 (99.5%) MARTHA and MEGA participants, respectively. Finally, the MARTHA *prospective sample* was composed of 839 individuals including 159 recurrences, the extended MARTHA *ambispective sample* included 1,504 among which 565 recurrences were observed and the MEGA sample was composed of 1,248 individuals including 428 recurrences. A detailed flow chart of the MARTHA sub-samples is presented in Supplementary Figure S1.

### Modelling strategy

Association of *ABO* blood groups with first recurrence was tested assuming additive effects of *ABO* tagging polymorphims, using the O1 group as a reference. Analyses were adjusted for sex, provoked status of the first VT (corresponding to the presence of a risk factor which temporarily promotes VT such as pregnancy or surgery), age at first VT, type of the first VT (DVT or PE) and the first 4 principal components derived from the GWAS genotypic data, in accordance with the literature.[7]

Finally, *ABO* association parameters from the MARTHA *ambispective* and MEGA analyses were meta-analysed using a fixed-effects model (Mantel-Haenszel methodology) to highlight the observed trends.[34]

## RESULTS

### Population characteristics

The main characteristics of the MARTHA *prospective* and extended *ambispective* samples are shown in Table 1. MARTHA patients were included in the study at approximately 47 years old with a mean age at the first VT around 41. Patients were included in average 6 years after their first VT. The distribution of the age at enrolment and the delay between enrolment and the first VT are provided in Supplementary Figures S2 & S3. For approximately 80% of patients, the first VT was DVT and in two-thirds of individuals the first VT was provoked. One-third of patients were male, with a higher proportion of men in those with recurrences. On average, patients were followed for 9 years and this time was longer for patients without recurrence in both samples, since follow-up ends at the occurrence of a recurrence. Regarding *ABO* blood groups, O1 was the most frequent (∼50%) followed by A1 (∼33%), B (∼9%), A2 (∼6%) and finally O2 (∼2%).

**Table 1:** Description of the main characteristics in the *prospective* and *ambispective* MARTHA samples.

| Variables | MARTHA *Prospective* sample |  |  | MARTHA *Ambispective* sample |  |  |
| --- | --- | --- | --- | --- | --- | --- |
|  | Total<br>N=839 | Recurrences<br>N=159 | Non-recurrences<br>N=680 | Total<br>N=1,504 | Recurrences<br>N=565 | Non-recurrences<br>N=939 |
|  | N (%) | N (%) | N (%) | N (%) | N (%) | N (%) |
| **Gender** |  |  |  |  |  |  |
| Men | 275 (32.8%) | 63 (39.6%) | 212 (31.2%) | 509 (33.8%) | 232 (41.1%) | 277 (29.5%) |
| **Age at inclusion** (mean ± SD) | 45.2 ± 14.9 | 44.0 ± 13.9 | 45.5 ± 15.4 | 47.1 ± 15.4 | 50.0 ± 14.7 | 45.3 ± 15.5 |
| **Age at the first VT** (mean ± SD) | 42.3 ± 15.5 | 41.5 ± 14.4 | 42.5 ± 15.8 | 41.0 ± 15.7 | 40.5 ± 15.1 | 41.3 ± 16.0 |
| **Delay between inclusion and first VT**<br>(In years, mean ± SD) | 2.9 ± 6.2 | 2.5 ± 5.3 | 3.0 ± 6.4 | 6.1 ± 9.85 | 9.5 ± 11.3 | 4.0 ± 8.2 |
| **Type of the first VT** |  |  |  |  |  |  |
| DVT | 653 (77.8%) | 122 (76.7%) | 531 (78.1%) | 1,189 (79.1%) | 454 (80.4%) | 735 (78.3%) |
| **Characteristic of the first VT** |  |  |  |  |  |  |
| Provoked | 544 (64.8%) | 93 (58.5%) | 451 (66.3%) | 993 (66.0%) | 368 (65.1%) | 625 (66.6%) |
| **Age at the collection of information on recurrence** (mean ± SD)* | 54.9 ± 15.2 | 54.9 ± 13.9 | 54.9 ± 15.5 | 52.5 ± 15.6 | 43.0 ± 14.2 | 52.1 ± 16.4 |
| **Delay of follow-up in years\*\***<br>(In years, mean ± SD) | 8.8 ± 5.5 | 6.4 ± 5.3 | 9.4 ± 5.4 | 9.7 ± 9.6 | 7.9 ± 8.9 | 10.8 ± 9.8 |
| **ABO haplotypes** |  |  |  |  |  |  |
| A1 | 32.8% | 37.4% | 31.7% | 33.5% | 35.8% | 32.2% |
| A2 | 5.5% | 5.3% | 5.5% | 5.9% | 6.9% | 5.3% |
| O1 | 51.0% | 47.5% | 51.8% | 49.8% | 46.9% | 51.5% |
| O2 | 1.7% | 1.6% | 1.7% | 1.5% | 1.5% | 1.5% |
| B | 9.1% | 8.2% | 9.3% | 9.3% | 8.9% | 9.5% |

\*Since inclusion in the prospective sample, since the first VT in the ambispective sample
\*\*Refers to the recontact for Cases 1 & 2 and inclusion for Cases 3 & 4 (see Fig1)

A description of the principal characteristics of the MEGA participants is provided in Table 2. The main differences with MARTHA sample are a higher proportion of men (49%), a higher age at first VT (∼48yrs), a lower rate of DVT (61%) and a shorter (∼5yrs) follow-up. The delay between inclusion and the first VT is not presented as only incident VT cases were recruited.

**Table 2.** Description of the main characteristics in the MEGA sample.

| Variables | MEGA sample |  |  |
| --- | --- | --- | --- |
|  | Total<br>N=1,248 | Recurrences<br>N=428 | Non-recurrences<br>N=820 |
|  | N (%) | N (%) | N (%) |
| **Gender** |  |  |  |
| Men | 661 (49.0%) | 272 (63.6%) | 389 (47.4%) |
| **Age at the first VT (mean ± SD)** | 48.0 ± 12.8 | 49.8 ± 12.8 | 47.1 ± 12.8 |
| **Type of the first VT** |  |  |  |
| DVT | 763 (61.1%) | 270 (63.1%) | 493 (60.1%) |
| **Characteristic of the first VT** |  |  |  |
| Provoked | 847 (67.9%) | 237 (55.4%) | 610 (74.4%) |
| **Delay of follow-up since inclusion**<br>(In years, mean ± SD) | 5.2 ± 2.9 | 3.1 ± 2.2 | 6.3 ± 2.7 |
| **ABO haplotypes** |  |  |  |
| A1 | 28.9% | 31.8% | 27.4% |
| A2 | 7.0% | 7.7% | 6.6% |
| O1 | 53.4% | 50.9% | 54.7% |
| O2 | 1.8% | 1.5% | 2.0% |
| B | 8.9% | 8.1% | 9.3% |

The sample used to estimate the risk of death in MARTHA was composed of 1,380 patients among whom 73 deaths were observed (Supplementary Table S1). The mean time of follow-up according to the last known vital status was around 12 years and other characteristics were similar to the MARTHA *ambispective* sample.

### Risk of death estimation

We estimated the risk of death in MARTHA with a delayed-entry Cox model (Supplementary Figure S4). The explanatory variables of this model were sex, provoked character of the first VT, age at the first VT and the first four principal components of the population stratification. Men had an HR (95% Confidence Interval) for death of 1.44 (0.87-2.40) whereas the provoked character of the first VT (HR=0.42 (0.24-0.74)) and a higher age of first VT (HR=0.98 (0.96-1.00) per year) were associated with reduced risk of death.

Using this model, we estimated the survival probability of patients up to the time at which information on their possible VT recurrence was collected. As described in *Methods* section, weights were based on survival probabilities and their range varied between 0.9 and 2.3 (Supplementary Figure S5).

### Clinical variables and VT recurrence risk

As a first step, we assessed the association of non-time dependent clinical variables on the risk of first VT recurrence in the MARTHA *prospective* and *ambispective* samples (Table 3). In the *prospective* analysis of 839 subjects including 159 recurrences, male sex was associated with an increased risk of VT recurrence (HR=1.47 (1.03-2.09)). Other variables were not significantly associated with recurrence, but a trend was observed for the provoked character of the first VT that tends to be protective (HR=0.69 (0.48-1.00)).

**Table 3.** Association of clinical variables and ABO haplotypes with VT recurrence in MARTHA (*prospective* and *ambispective*) and MEGA.

| Variables | MARTHA *Prospective* |  | MARTHA *Ambispective* |  | MEGA |  | Meta-analysis<br>MARTHA *Ambispective*<br>& MEGA |  |
| --- | --- | --- | --- | --- | --- | --- | --- | --- |
|  | N=839 |  | N=1,504 |  | N=1,248 |  |  |  |
|  | Nb recurrences=159 |  | Nb recurrences=565 |  | Nb recurrences=428 |  |  |  |
|  | HR (95% CI) | P | HR (95% CI) | P | HR (95% CI) | P | HR (95% CI) | P |
| **Gender** |  |  |  |  |  |  |  |  |
| Men | 1.47 (1.03-2.09) | 0.034 | 1.65 (1.36-2.01) | 4.0x10<sup>-7</sup> | 1.81 (1.46-2.25) | 5.9x10<sup>-8</sup> | 1.72 (1.47-2.01) | 3.0x10<sup>-12</sup> |
| **Age at the first VT**<br>(10 years increase) | 0.91 (0.81-1.02) | 0.105 | 1.08 (1.02-1.15) | 0.020 | 0.99 (0.92-1.07) | 0.810 | 1.05 (0.99-1.11) | 0.107 |
| **Type of the first VT** |  |  |  |  |  |  |  |  |
| DVT | 0.85 (0.60-1.21) | 0.368 | 1.17 (0.96-1.42) | 0.140 | 1.15 (0.95-1.40) | 0.160 | 1.16 (1.01-1.33) | 0.036 |
| **Characteristic of the first VT** |  |  |  |  |  |  |  |  |
| Provoked | 0.69 (0.48-1.00) | 0.059 | 0.99 (0.80-1.23) | 0.920 | 0.61 (0.49-0.76) | 6.7x10<sup>-6</sup> | 0.78 (0.67-0.91) | 1.2x10<sup>-3</sup> |
| **ABO haplotypes** |  |  |  |  |  |  |  |  |
| A1 | 1.32 (1.02-1.70) | 0.035 | 1.15 (1.00-1.32) | 0.045 | 1.21 (1.03-1.42) | 0.018 | 1.18 (1.05-1.33) | 4.2x10<sup>-3</sup> |
| A2 | 1.13 (0.68-1.88) | 0.644 | 1.27 (0.98-1.64) | 0.061 | 1.11 (0.86-1.43) | 0.409 | 1.19 (1.00-1.42) | 0.062 |
| O1 | Reference |  | Reference |  | Reference |  | Reference |  |
| O2 | 1.05 (0.47-2.35) | 0.896 |  |  | 0.86 (0.50-1.49) | 0.584 | 1.03 (0.70-1.52) | 0.880 |
| B | 1.00 (0.64-1.57) | 0.998 | 1.02 (0.82-1.27) | 0.874 | 1.00 (0.78-1.29) | 0.987 | 1.01 (0.85-1.21) | 0.900 |

*HR: Hazard Ratio*
*CI: Confidence Interval*

The analysis of the same variables performed in the extended *ambispective* sample allows to refine some of these observations with a higher power. Male sex was still associated with a higher risk of recurrence HR=1.65 (1.36-2.01); and older age at first VT appeared as deleterious (HR=1.08 (1.02-1.15) for a 10 years increase). Conversely, we did not find any trend for the provoked status of the first VT (HR=0.99 (0.80-1.23)).

In MEGA, male sex was also associated with an increased risk of recurrence (HR=1.81) but older age was not. Besides, the provoked status of the first VT was significantly associated with a decreased risk of recurrence (HR=0.61), as initially observed in the MARTHA *prospective* analysis but not confirmed in the *ambispective* analysis.

Lastly, even if the type of first VT (DVT vs PE) was not significantly associated with VT recurrence in neither of the two studies, the same trend for a higher risk of recurrence associated with DVT was observed in the MARTHA *ambispective* (HR=1.17) and the MEGA (HR=1.15) samples. The meta-analysis of these two HRs yielded a combined HR of 1.16.

### ABO blood groups

In the MARTHA *prospective* sample, we observed a significant association of A1 blood group compared with O1 on the risk of first VT recurrence (HR=1.32 (1.02-1.70); p=0.035) which was confirmed in the analysis of the *ambispective* sample (HR=1.15 (1.00-1.32); p=0.045) (Table 3). The same trend was observed for the A2 group but did not reach statistical significance (HR=1.27 (0.98-1.64); p=0.061 in the *ambispective* analysis). In MEGA, only A1 was significantly associated with a higher risk of VT recurrence (HR=1.21; p=0.018). No evidence for association with VT recurrence was observed for B and O2 groups in either MARTHA or MEGA.

Finally, based on the meta-analysis of the results observed in the MARTHA *ambispective* and MEGA samples, the risk of VT recurrence associated with *ABO* blood groups compared with O1 were HR=1.18 (p=4.2×10^−3^), HR=1.19 (p=0.06), HR=1.01 (p=0.90) and HR=1.03 (p=0.88) for A1, A2, B and O2, respectively.

## DISCUSSION

The starting point of this work was to investigate the risk of VT recurrence associated with *ABO* blood groups in two large cohorts of VT patients, MARTHA and MEGA, the former being built upon an ambispective design.

To achieve this goal, we had first to propose a novel modelling approach to analyse non-time dependent risk factors (such as genetic polymorphisms) of an event which could have occurred in patients before their inclusion in the study. This modelling was mandatory to maximize the power of the MARTHA study as about 70% of first VT recurrences in MARTHA occurred in patients before their inclusion in the study. The proposed modelling relies on a weighted Cox model where the use of weights allows to limit the selection bias associated with the use of pre-inclusion events and thus to gain statistical power by jointly analysing pre- and post-inclusion recurrent events.

This method differs from the weighting approach for repeated events proposed to deal with event-dependent sampling.[35] Indeed, the inclusion in MARTHA depends on an event, the first VT, which is not the outcome of interest; the first VT defines the beginning of the period at risk for the recurrence. Our weighting approach handles potential bias due to mortality until the time of data collection for the recurrence.

Our proposed weighted estimation approach is unbiased if the weights are well-specified which means that the Cox model for death is correct. In this work, the death model has two main limitations. First, as the information on VT recurrence was often missing for subjects who died, it was not possible to include VT recurrence (and other possible unknown variables) as a risk factor in our model for death risk. Second, we assumed the proportionality of the risk of death and did not account for the calendar time that could modify either the baseline risk of death or the risk factors for death. Moreover, as the number of death during the follow-up in MARTHA is quite small, a Monte Carlo analysis was performed to evaluate the sensitivity of the results to the uncertainty on the weights (description is available in the Supplement). Despite some slight variability in HRs’ estimates (especially for O2 group), the overall results remain unchanged.

In this work, we were interested in the association of ABO blood groups with the risk of first VT recurrence and not with the risk of multiple VT recurrences. Indeed, at inclusion in MARTHA, only detailed information on first VT and possibly first recurrence was collected, preventing us from investigating the association with multiple recurrences. Besides, the analysis of such multiple events would require more complex modeling that would take into account the correlation of repeated events.[36]

The analysis of the MARTHA and MEGA studies, totaling 2,752 VT patients including 993 recurrences, revealed that the A1 and A2 blood groups were both associated with increased risk of VT recurrence, HR ∼1.20 for both, compared with O1. Note that, likely because of the modest frequency of the A2 blood group (∼5%), the association was only marginally significant (p=0.06). Some studies have already investigated the association of *ABO* blood groups with the risk of VT recurrence[11–14], but often with a moderate sample size or using serological *ABO* phenotypes whereas we here used genetically defined *ABO* blood groups which has been shown to be more efficient to capture the effect of *ABO* on VT risk.[15] Our results are consistent with those showing a higher risk of recurrence in non-OO patients.[12–14] However, they are discordant with the study of *Baudouy et al*. who found a higher risk of VT recurrence in B blood group patients[11], while no association (HR=1.01, p=0.90) was observed in our work. Beyond the rather modest size of their study (N=100) and the analysis of serological *ABO* phenotypes, the work of *Baudouy et al*. focused on patients with PE as first VT. As a consequence, we further stratified the association analysis of *ABO* blood groups with recurrence according to the type of the first event (DVT or PE) but did not observe any evidence for specific sub-group *ABO* effects (Supplementary Table S2).

As the proposed *ambispective* modelling is only valid for analysing non-time dependent variables, we could not adjust the *ABO* blood group’s effect on biological variables that have only been measured at the time of inclusion, such as von Willebrand Factor (vWF). Adjusting for vWF plasma levels would have allowed us to determine whether the effect of *ABO* on VT recurrence is mediated by vWF. This is however unlikely as the observed pattern of association of *ABO* blood groups with recurrence does not match the known associations between *ABO* blood groups and vWF plasma levels.[15] Furthermore, we could not adjust our analysis for the familial history of VT as the available information in MARTHA refers to the presence of a history at the time of inclusion and many recurrences arose earlier.

Nevertheless, our modelling enabled us to assess the impact on the risk of VT recurrence of several clinical variables that are fixed after the first VT event such age at first VT and the type of first VT (DVT vs PE; provoked vs unprovoked). Consistent in MARTHA and in MEGA were the associations of male sex and DVT as first VT with an increased risk of first VT recurrence, confirming previous observations.[7,37] However, we did not observe consistent results with respect to age at first VT nor with the provoked status of the first VT. The different trends observed can be due to the different design and sample selection between MARTHA and MEGA. Indeed, participants in MARTHA were included for at least one previous VT which may have occurred more than fifty years before their inclusion (Mean=6years; Standard Error=10years) whereas in MEGA, patients were recruited at the time of their first VT. Besides, the definition of the provoked character slightly differs between MARTHA and MEGA (Supplementary Table S3). We also observed some differences between MARTHA *prospective* and *ambispective* that might be due to the calendar time which has not been taken into account in our work. In MARTHA ∼25% of the patients had their first VT between the 1940s and the start of the study (1994); these VT were provoked in 80% of cases. We are aware that the differences in the management, prevention and identification of VT events at that period may have masked the association between the provoked character of the first event and VT recurrence in the *ambispective* analysis. However, we feel that such differences may have modest impact when one is interested in genetic factors as illustrated here with the consistent patterns observed for *ABO* blood groups in both MARTHA *prospective* and *ambispective* samples as well as in MEGA.

In conclusion, this study demonstrated that both A1 and A2 blood groups are associated with increased risk of VT recurrence. This finding was made possible thanks to a weighting approach to study non-time dependent risk factors while integrating not only post-inclusion events but also those that occurred before inclusion. This modelling finds an immediate field of application to genetic association studies for time-to-events in cohorts where follow-up information for deaths is available and events before inclusion are collected.

## Data Availability

All data produced in the present study are available upon reasonable request to the authors.

## Acknowledgments

GM benefited from the EUR DPH, a PhD program supported within the framework of the PIA3 (Investment for the future). Project reference 17-EURE-0019.

This project was carried out in the framework of the INSERM GOLD Cross-Cutting program (P-EM, D-AT). Statistical analyses benefited from the CBiB computing centre of the University of Bordeaux.

## Ethics approval and consent to participate

Written informed consent was obtained from all MARTHA and MEGA participants. The MARTHA study was approved by the local Ethics Committee at La Timone Hospital (Marseille, France). The MEGA study was approved by the Medical Ethics Committee of the Leiden University Medical Center, the Netherlands.

## Contributions

GM performed statistical analyses and wrote the first draft of the paper. HJG and D-AT supervised the statistical analyses and revised the paper. LG, AvHV, MI-K, MB, J-FD, FR and P-EM participated to data collection. FR, P-EM and D-AT. designed the study. All authors read and approved the final manuscript.

