## Supplementary Materials for "Modelling of time-to-events in an ambispective study: illustration with the analysis of *ABO* blood groups on venous thrombosis recurrence"

### CONTENTS:

Supplementary Figures 1-6 2

Supplementary Tables 1-3 5

Supplementary Text 8

Supplementary Figure S1. Flow chart of the MARTHA sub-samples

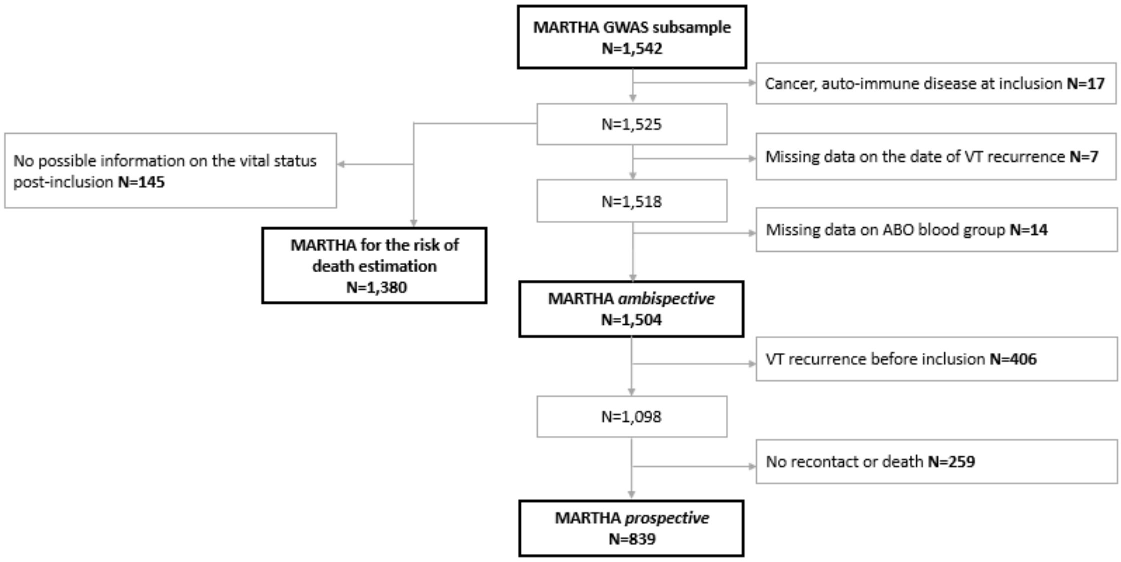


Supplementary Figure S2. Distribution of the age at enrolment in MARTHA participants (N=1,504)

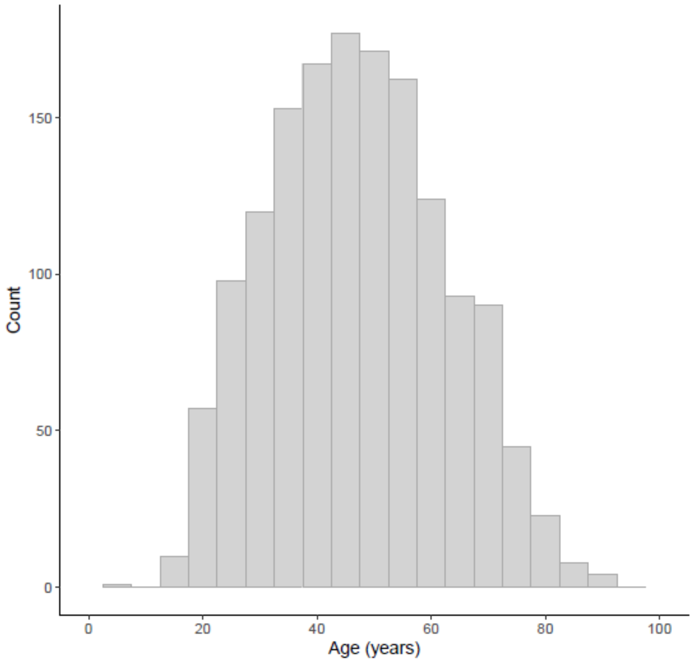

Supplementary Figure S3. Distribution of the delay between enrolment and the first VT in MARTHA participants (N=1,504)

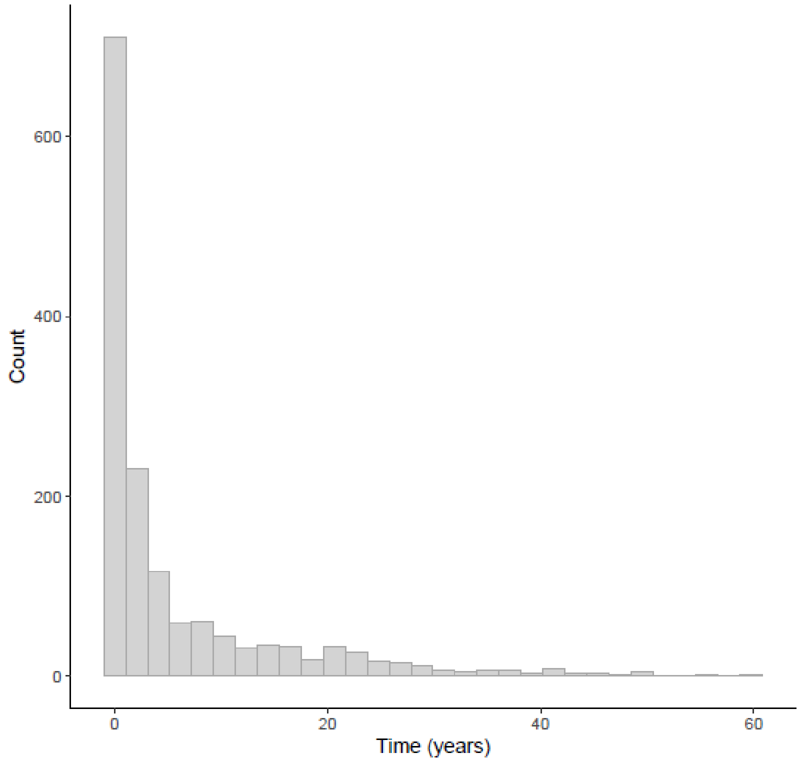


Supplementary Figure S4. Kaplan Meier plot of the survival probability in MARTHA participants with an available follow up (N=1,380 including 73 deaths)

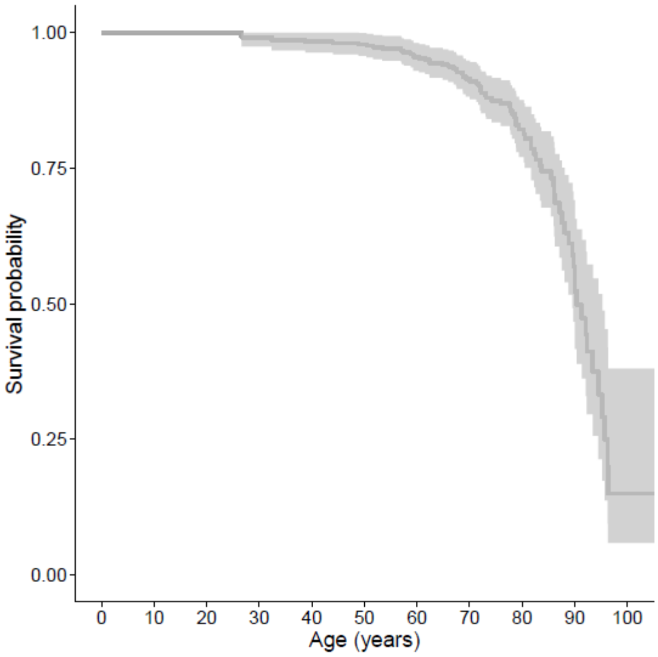

Supplementary Figure S5. Distribution of the estimated weights for the MARTHA participants (N=1,504)

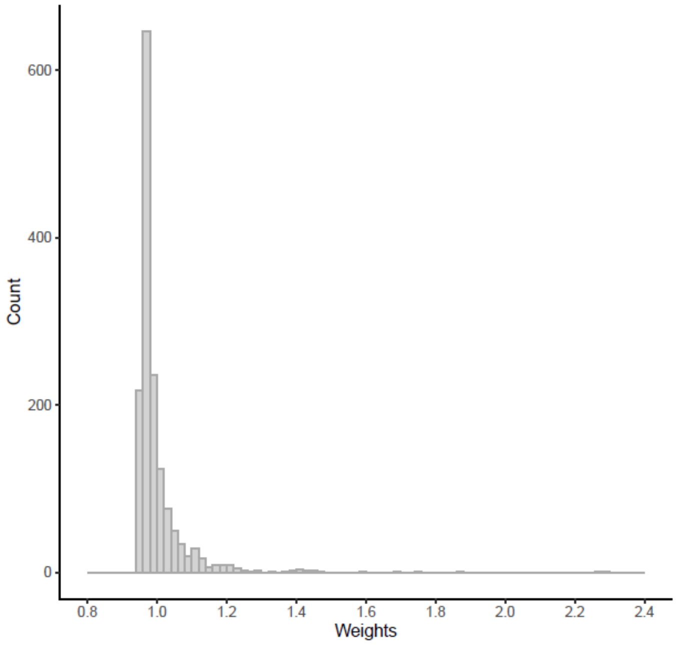


Supplementary Figure S6. Sensitivity of the association of *ABO* blood groups with recurrence according to the weights estimation in MARTHA (N=1,504)

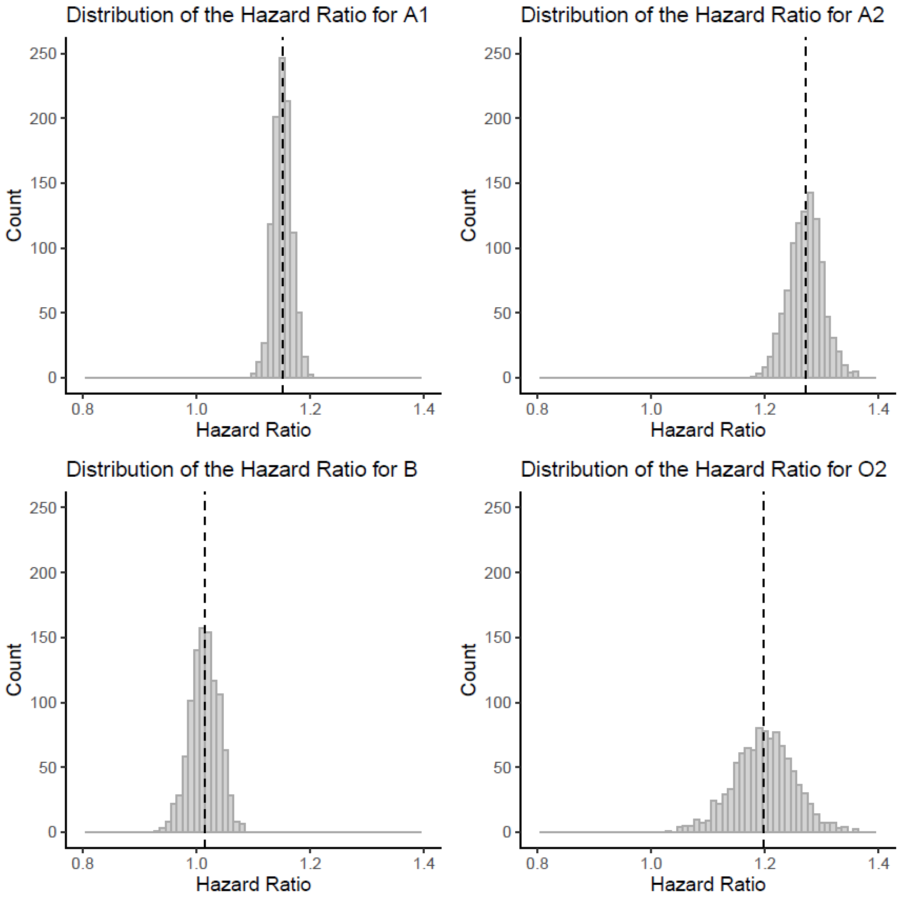


Note: The 4 panels show the distribution of the Hazard Ratio in the Monte Carlo resampling analysis (See Supplementary Text). The dashed line corresponds to the estimated value in the initial model

Supplementary Table S1. Description of the MARTHA sample for the death risk estimation

| **Variables** | **Total N=1,380** |
|---|---|
| | **N (%)** |
| **Gender** | |
| Men | 468 (33.9%) |
| **Age at inclusion** (mean ± Standard Deviation (SD)) | 47.1 ± 15.3 |
| **Age at the first VT** (mean ± SD) | 41.3 ± 15.7 |
| **Delay between inclusion and first VT** (In years, mean ± SD) | 5.8 ± 9.6 |
| **Type of the first VT** | |
| DVT only | 1,087 (78.8%) |
| **Characteristic of the first VT** | |
| Provoked | 911 (66.0%) |
| **Delay of follow-up in years*** (In years, mean ± SD) | 11.8 ± 5.3 |

*According to the death event

Supplementary Table S2. Association of *ABO* haplotypes with first VT recurrence in MARTHA *ambispective* and MEGA stratified on the type of the first VT

| **Variables** | **MARTHA *Ambispective*** **N=1,504** **Nb recurrences=565** | | **MEGA** **N=1,248** **Nb recurrences=428** | | **Meta-Analysis Fixed-effects** | |
|---|---|---|---|---|---|---|
| | **HR (95% CI)** | **P** | **HR (95% CI)** | **P** | **HR (95% CI)** | **P** |
| **ABO haplotypes – PE as first VT** | **N=315 ; 111 recurrences** | | **N=485 ; 158 recurrences** | | | |
| A1 | 1.10 (0.82-1.48) | 0.536 | 1.38 (1.05-1.82) | 0.020 | 1.24 (1.02-1.51) | 0.029 |
| A2 | 1.87 (1.04-3.37) | 0.039 | 0.79 (0.49-1.27) | 0.329 | 1.11 (0.80-1.55) | 0.554 |
| O1 | Reference | | Reference | | Reference | |
| O2 | 0.65 (0.13-3.24) | 0.600 | 0.70 (0.28-1.72) | 0.434 | 0.69 (0.36-1.32) | 0.250 |
| B | 0.85 (0.47-1.53) | 0.574 | 0.83 (0.51-1.36) | 0.447 | 0.84 (0.59-1.20) | 0.318 |
| **ABO haplotypes – DVT as first VT** | **N=1,189 ; 454 recurrences** | | **N=763 ; 270 recurrences** | | | |
| A1 | 1.16 (0.99-1.36) | 0.063 | 1.14 (0.94-1.39) | 0.211 | 1.15 (1.00-1.32) | 0.045 |
| A2 | 1.20 (0.91-1.58) | 0.180 | 1.29 (0.94-1.77) | 0.104 | 1.24 (1.00-1.54) | 0.059 |
| O1 | Reference | | Reference | | Reference | |
| O2 | 1.41 (0.85-2.35) | 0.180 | 0.94 (0.46-1.90) | 0.872 | 1.23 (0.75-2.01) | 0.422 |
| B | 1.06 (0.84-1.34) | 0.612 | 1.08 (0.79-1.48) | 0.637 | 1.07 (0.86-1.33) | 0.566 |

*HR: Hazard Ratio*

*CI: Confidence Interval*

Supplementary Table S3. Definition of the provoked character in MARTHA and MEGA

| MARTHA study | MEGA study |
|---|---|
| • Surgery within 3 months before VT | • Surgery within 3 months before VT |
| • Pregnancy/ puerperium within 3 months before VT | • Pregnancy/ puerperium within 3 months before VT |
| • Oral contraceptive use within 3 months before VT | • Hormone use at the time of VT, including: hormone replacement therapy and hormonal contraceptives |
| | • Plaster cast within 3 months before VT |
| • Immobilization for 7 days or more within 3 months before VT | • Immobility in bed, in hospital: Confinement to bed ≥ 3 days in hospital, confinement to bed ≥ 3 days at home, within 3 months before VT |
| • Long travel (by car >10 hours ; by plane > 5 hours) within 3 months before VT | • Prolonged travel >4 hours within 2 months before VT |
| • Trauma of the lower limb within 3 months before VT | • Leg injury in 3 months before VT |
| • Pneumonia in year before VT | • Pneumonia in year before VT |
| • Infection in year before VT (urinary tract infection, pyelonephritis, arthritis, bursitis, sinusitis, pulpitis, inflammation elsewhere, hepatitis A, B or C) | • Infection in year before VT (urinary tract infection, pyelonephritis, arthritis, bursitis, sinusitis, pulpitis, inflammation elsewhere, hepatitis A, B or C) |

Supplementary Text. Sensitivity analysis on the weights estimation for MARTHA participants

*Methods:* To investigate the variability of the weights estimated from the MARTHA study and their impact on the weighted Cox model, we used a Monte Carlo method. From the death risk model, we estimated the survival function $\hat{S}(t_i|Z_i) = \exp(-\hat{A}(t_i|Z_i))$ of each individual $i$ up to the time $t_i$ (which corresponds to the time of collection of the information on VT recurrence). Assuming that the cumulative risk $\hat{A}(t_i|Z_i)$ follows a normal distribution, for each individual we randomly draw 1,000 values of the his/her cumulative risk from the distributions $N(\hat{A}(t_i|Z_i), SE(\hat{A}(t_i|Z_i)))$ and computed the corresponding survival probabilities $\hat{S}_k(t_i|Z_i) = \exp(-\hat{A}_k(t_i|Z_i))$ to obtain the set of individual weights $w_{ik}$ for k=1,…,1000. Then 1,000 weighted Cox model for the VT recurrence were estimated.

*Results*: The distributions of the HR for the ABO blood groups from the 1,000 models for VT recurrence are shown in Supplementary Figure 6 where the value of the HR estimated in the initial model is presented as a dashed line. The empirical distributions are well centred at the initial estimated HRs and, for both A1 and A2, all estimated HRs are above 1 supporting our conclusions.
